# Development of a prediction model for 30-day COVID-19 hospitalization and death in a national cohort of Veterans Health Administration patients – March 2022 - April 2023

**DOI:** 10.1101/2023.11.17.23298653

**Authors:** David P. Bui, Kristina L. Bajema, Yuan Huang, Lei Yan, Yuli Li, Rajeevan Nallakkandi, Kristin Berry, Mazhgan Rowneki, Stephanie Argraves, Denise Hynes, Grant Huang, Mihaela Aslan, George N. Ioannou

## Abstract

**Objective:** Develop models to predict 30-day COVID-19 hospitalization and death in the Omicron era for clinical and research applications.

**Material and Methods:** We used comprehensive electronic health records from a national cohort of patients in the Veterans Health Administration (VHA) who tested positive for SARS-CoV-2 between March 1, 2022, and March 31, 2023. Full models incorporated 84 predictors, including demographics, comorbidities, and receipt of COVID-19 vaccinations and anti-SARS-CoV-2 treatments. Parsimonious models included 19 predictors. We created models for 30-day hospitalization or death, 30-day hospitalization, and 30-day all-cause mortality. We used the Super Learner ensemble machine learning algorithm to fit prediction models. Model performance was assessed with the area under the receiver operating characteristic curve (AUC), Brier scores, and calibration intercepts and slopes in a 20% holdout dataset.

**Results:** Models were trained and tested on 198,174 patients, of whom 8% were hospitalized or died within 30 days of testing positive. AUCs for the full models ranged from 0.80 (hospitalization) to 0.91 (death). Brier scores were close to 0, with the lowest error in the mortality model (Brier score: 0.01). All three models were well calibrated with calibration intercepts <0.23 and slopes <1.05. Parsimonious models performed comparably to full models.

**Discussion:** These models may be used for risk stratification to inform COVID-19 treatment and to identify high-risk patients for inclusion in clinical trials.

**Conclusions:** We developed prediction models that accurately estimate COVID-19 hospitalization and mortality risk following emergence of the Omicron variant and in the setting of COVID-19 vaccinations and antiviral treatments.

## INTRODUCTION

Early in the COVID-19 pandemic, many prognostic prediction models were developed to identify persons infected with SARS-CoV-2 who could be at high risk for adverse outcomes (1–3). These models informed implementation of risk reduction measures, clinical care, and resource allocation (4). Despite reductions in mortality and hospitalization rates, COVID-19 remains a leading cause of death in the United States and accounts for thousands of hospitalizations every week nationally (5). COVID-19 risk prediction models may continue to inform patient care and policy decisions, particularly as effective vaccines and antiviral treatments have become available (6,7). Such models may also help identify high-risk patients for facilitating future clinical trials of new pharmacotherapies.

Currently available COVID-19 risk prediction models have important limitations. First, most models were developed early in the pandemic (8–13) and do not account for current circulating viral strains, population immunity from previous infection or vaccination, and treatments. This is particularly important as risk models are sensitive to population contextual factors under which they are developed. Model performance, especially calibration, may diminish over time as context (e.g., population immunity, treatment availability) changes (14,15). Moreover, most models were developed among acutely ill patients receiving care in the hospital (13,16–18) and emergency department settings (19), focusing predominantly on mortality and severe outcomes, and may not be applicable to or perform well in a broader population, particularly as mortality rates have declined. Most COVID-19 risk models have largely relied on parametric regression models with strong assumptions of linearity (e.g., logistic regression) which may not be met for many predictors. Further, most available models rarely leverage ensemble methods which can utilize parametric and nonparametric modeling approaches to hedge against model misspecification and have been shown to improve predictive performance in diverse applications (20–22).

We report the development, assessment, and internal validation of multivariable risk prediction models for 30-day hospitalization and death among patients with COVID-19, which use more recent data to reflect the current context of COVID-19 and leverage an ensemble modeling approach. These models were developed on a large national source population of United States Veterans enrolled in Veterans Health Administration (VHA) care using detailed covariates captured in electronic health records (EHR).

## MATERIAL AND METHODS

### Data sources, Participants, and Sample Size

We used the Department of Veterans Affairs (VA) COVID-19 Shared Data Resource (CSDR) to identify patients with COVID-19. The CSDR includes data on all VHA patients who had laboratory-confirmed SARS-CoV-2 tests performed in the VHA as well as tests performed outside the VHA and documented in clinical notes. In addition to CSDR, we used the VA Corporate Data Warehouse, a repository of VA comprehensive electronic health records, which includes a broad range of demographic (e.g., age, sex, race), geographic, and clinical variables (e.g., vaccination, comorbidity indicators) in our prediction models. Hospitalization and mortality outcomes obtained from CSDR, and hospitalizations were further supplemented with claims data for purchased care provided outside of the VHA through the VHA Integrated Veteran Care Program and with Fee-for-Service claims from the Centers for Medicare & Medicaid Services (CMS). CMS data were also used to supplement data on COVID-19 vaccinations.

For model development and validation, we selected all patients aged 18 years and older with a first positive SARS-CoV-2 test in CSDR between March 1, 2022, and March 31, 2023. Patients from all VHA facilities across 50 states and territories were eligible for inclusion. We limited the cohort to patients obtaining VHA care, define as having a VHA primary care encounter in the 18 months preceding the positive test to minimize missing data and improve ascertainment of key variables.

### Outcomes

We developed separate models to predict three outcomes: 1) 30-day hospitalization or death (composite outcome model), 2) 30-day hospitalization (hospitalization model), and 3) 30-day all-cause mortality (mortality model). Hospitalizations and deaths were counted if they occurred within 30 days from the first test-positive date for each patient.

### Predictors

We reviewed predictors available in the EHR and selected covariates for model development based on prior research and clinical knowledge (6,24–26). We selected 84 predictors, including demographic variables (e.g., age, sex, race), geographic variables (e.g., region of residence, rurality), body mass index, indicators for comorbidities, COVID-19 vaccination status, and receipt of outpatient anti-SARS-CoV-2 treatments, including nirmatrelvir-ritonavir, molnupiravir, and monoclonal antibodies. COVID-19 treatments were considered in the list of candidate predictors as studies have shown including treatments known to affect outcomes can enhance predictive performance and precision in prognostic models (27,28); inclusion of post-infection treatment further flexibly allows prediction of pre-treatment and on-treatment risk. We included both individual comorbidity indicators (e.g., type 1 diabetes, type 2 diabetes, diabetes with complications) as well as composite indicators of related comorbidities (e.g., any diabetes). We also included three index measures of vulnerability and frailty, including the Area Deprivation Index (ADI) (29) the Charlson Comorbidity Index (CCI) (30), and the Care Assessment Needs (CAN) score (31), an automated EHR-based risk score developed at the VHA to predict 1-year mortality (32). See Supplement S1 for full list of predictors.

### Training and holdout test datasets

We used a random 80/20 data split of our full cohort to create a training and a holdout dataset, respectively. Sampling was done at the VHA facility level such that all patients from a given station were either allocated to the training dataset or test dataset but not both. This data partition approach ensured our models were developed on a population that was geographically different from the population in which we tested model performance.

### Data Pre-Processing and Missing Data

We pre-processed the training dataset to ensure categorical variables were properly modeled and missing data was imputed. Missing data was rare, with just 3,971 (2%) of our cohort missing one or more variables; the CAN score was the most frequently missing variable and was missing for 2,883 (1.4%) of patients. Categorical predictors were one-hot coded by creating a series of binary ‘dummy’ indicator variables for each factor level with a reference level omitted; most prevalent levels served as the referent. Missing continuous covariates were imputed with observed medians, and missing categorical covariates were imputed with observed modes. We created covariate-specific missing indicators (0/1) to flag observations with missing and imputed covariate values to allow covariate missingness to be an outcome predictor, which will allow future prediction with missing data (33,34).

### Statistical analysis methods: model development

We used the Super Learner (SL) algorithm to develop a risk prediction model for each specified outcome (20). SL is an ensemble machine learning algorithm that leverages cross-validation to find optimal weighted combinations of a pre-specified ensemble of models and algorithms (i.e., learners) to improve prediction. The SL algorithm requires minimal assumptions, is flexible enough to include both parametric and nonparametric learners, and mitigates the risk of overfitting in high-dimensional applications through cross-validation making it an ideal algorithm for complex prediction problems (21,22,34). The SL algorithm has been applied in numerous prediction (35) and causal inference studies and shown to perform as well as or better than the best performing individual learner, and is especially well-suited for rare outcomes (21,36). We used the ‘SuperLearner’ R package (version 2.0-28.1) to build our SL models.

In total, our final ensembles included a library of 10 distinct learners, including general linear models, penalized regressions (LASSO and ridge regression LASSO and ridge regression), and machine learning algorithms such as generalized additive models, multivariate adaptive regression splines, neural networks, random forest, and XGBoost (see Supplement S2 for learners and hyperparameters). For machine learning algorithms included in the ensemble, we used the ‘caret’ package (version 6.0-94) to perform a series of cross-validated grid searches of hyperparameters in a random 50% sample of the training dataset (to reduce computation time) and used the set of parameters that maximized the cross-validated area under the receiver operating characteristic curve (AUC) for the composite hospitalization or death outcome model (37).

To fit ensemble models, we used a 5-fold cross-validation scheme stratified on outcome to generate cross-validated predictions from each included learner, which were then used in a meta-learner to obtain final ensemble model weights for prediction. Based on our effective sample size and guidance from Phillips et al. (22), we chose 5-fold cross-validation for fitting our SL models to minimize computation time and bias in cross-validated risk estimates. We used the non-negative binomial likelihood maximization meta learner in the ‘SuperLearner’ package which has been shown to perform well for SL classification models (36) (see Supplement S3 for full SL model specifications).

### Assessing ensemble model performance and variable importance

For each fitted outcome model, we assessed model performance in two ways. First, we conducted 5-fold cross-validation of each ensemble model with the training dataset to estimate the ensemble model’s performance in unseen data with average AUC (a measure of discrimination, with 1 indicating perfect discrimination) as the target performance metric. Second, we assessed each model’s performance in the holdout dataset and reported each model’s AUC, Brier score (a measure of accuracy, with 0 indicating perfect accuracy), and Cox’s calibration intercept and slope (an intercept of 0 and slope of 1 indicating perfect calibration); no model updating or recalibration was conducted (38,39). Model calibration was also visually assessed in the holdout test dataset using local regression-smoothed calibration curves. We further estimated calibration intercepts and slopes for each model across various subgroups in the holdout dataset to assess local model calibration by age, race, ethnicity, region, and CAN score (39).

To compare our models’ performance against existing risk stratification scores, we estimated AUC, Brier scores, and calibration intercepts and slopes for the CAN score and the VACO Index, a publicly available risk score for 30-day COVID-19 mortality that was developed in the VHA population early in the pandemic (11,40). For the CAN score, we used Platt scaling to recalibrate the score to our COVID-19 outcomes in the training model dataset (39). The CAN score was modeled using restricted cubic splines with five knots and outcomes were predicted and assessed in the holdout dataset. We also calculated the absolute net reclassification index (NRI) for our full mortality model compared with CAN and VACO index to assess the improvement in our models’ ability to classify patients into lower and high risk groups (41). The absolute NRI measures the absolute percent of patients correctly reclassified in our developed model; greater positive NRIs suggest improvements in classification. Model performance metrics and NRI were estimated in the holdout test dataset.

To understand variable importance from our SL models, we calculated SHapley Additive exPlanations (SHAP) values from each model to understand the average marginal contribution of each variable to the predicted outcome probability (42). For computational efficiency, we estimated SHAP values from the individual learner assigned the most weight in each ensemble. SHAP values were estimated using the ‘shapviz’ R package (version 0.9.2). For visual clarity, we collapsed comorbidity indicators in the same domain (e.g., indicators for heart disease and heart failure were collapsed into ‘cardiovascular diseases’) and collapsed one-hot coded categorical variables (e.g., individual race indicators) and plot the top 15 feature domains in terms of largest average absolute SHAP value.

### Parsimonious model development

In addition to the full models, we developed and assessed a set of parsimonious models for each outcome which included the subset of most important predictors from our full models (the top 15 feature domains with highest SHAP values). To facilitate future model implementation in other healthcare systems, we omitted variables used exclusively within VHA, including the CAN score, facility complexity indicator, and residency at a Community Living Center (VA nursing home). These models were trained using the same learners and parameters used to fit the full ensemble models. We use the same performance metrics used to assess our full models and further calculate the absolute NRI for these parsimonious models compared with the full models. All analyses were conducted in R (version 4.3.1).

We followed the Transparent Reporting of a multivariable prediction model for Individual Prognosis or Diagnosis (TRIPOD) statement in preparing this manuscript (23). The study was approved by the VA Central Institutional Review Board.

## RESULTS

### Participants

We identified 198,174 VHA patients engaged in VHA care with a first positive COVID-19 test between March 1, 2022, and March 31, 2023; the cohort was predominantly male (174,110 [88%]), White (133,461 [67%]), non-Hispanic or Latino (166,670 [84%]), and with average age of 63 years (Table 1). Comorbid conditions were common, with 141,256 (71%) having cardiovascular disease, 91,144 (46%) having a mental health diagnosis (including bipolar disorder, depression, post-traumatic stress disorder, and schizophrenia), and 19,999 (10%) who were immunocompromised. Most (156,956 [79%]) were COVID-19 vaccinated. The patients in the training and holdout test datasets were similar on nearly all characteristics except race (SMD=0.18) and region (SMD=0.35). There were 15,037 (8%) persons in the cohort who were hospitalized or died within 30-days of testing positive, including 2,444 (1%) who died (Table 1).

**Table 1.** Cohort characteristics.

| Characteristic | Overall,<br>N = 198,174<sup>†</sup> | Training,<br>N = 158,748<sup>†</sup> | Holdout,<br>N = 39,426<sup>†</sup> | SMD |
| --- | --- | --- | --- | --- |
| **Age (years)** | 63.7 (15.51) | 63.6 (15.54) | 64.0 (15.38) | -0.026 |
| Unknown | 1 | 1 | 0 |  |
| **Male sex** | 174,110 (87.9%) | 139,175 (87.7%) | 34,935 (88.6%) | -0.029 |
| **Race** |  |  |  | 0.183 |
| American Indian/Alaska Native | 1,541 (0.8%) | 1,199 (0.8%) | 342 (0.9%) |  |
| Asian | 3,015 (1.5%) | 2,682 (1.7%) | 333 (0.8%) |  |
| Black or African American | 42,285 (21.3%) | 35,815 (22.6%) | 6,470 (16.4%) |  |
| Native Hawaiian/Pacific Islander | 1,938 (1.0%) | 1,610 (1.0%) | 328 (0.8%) |  |
| White | 133,461 (67.3%) | 105,233 (66.3%) | 28,228 (71.6%) |  |
| Unknown | 15,934 (8.0%) | 12,209 (7.7%) | 3,725 (9.4%) |  |
| **Ethnicity** |  |  |  | 0.009 |
| Not Hispanic or Latino | 166,670 (84.1%) | 133,613 (84.2%) | 33,057 (83.8%) |  |
| Hispanic or Latino | 18,765 (9.5%) | 14,978 (9.4%) | 3,787 (9.6%) |  |
| Unknown | 12,739 (6.4%) | 10,157 (6.4%) | 2,582 (6.5%) |  |
| **Smoking Status** |  |  |  | 0.050 |
| Never | 80,256 (40.5%) | 64,869 (40.9%) | 15,387 (39.0%) |  |
| Former | 80,316 (40.5%) | 64,216 (40.5%) | 16,100 (40.8%) |  |
| Current | 30,920 (15.6%) | 24,264 (15.3%) | 6,656 (16.9%) |  |
| Unknown | 6,682 (3.4%) | 5,399 (3.4%) | 1,283 (3.3%) |  |
| **Rurality** |  |  |  | 0.040 |
| Rural | 50,747 (25.9%) | 40,097 (25.5%) | 10,650 (27.3%) |  |
| Urban | 145,474 (74.1%) | 117,101 (74.5%) | 28,373 (72.7%) |  |
| Unknown | 1,953 | 1,550 | 403 |  |
| **Region** |  |  |  | 0.350 |
| South | 78,360 (39.5%) | 60,599 (38.2%) | 17,761 (45.0%) |  |
| West | 47,701 (24.1%) | 41,233 (26.0%) | 6,468 (16.4%) |  |
| Midwest | 36,781 (18.6%) | 26,593 (16.8%) | 10,188 (25.8%) |  |
| Northeast | 35,332 (17.8%) | 30,323 (19.1%) | 5,009 (12.7%) |  |
| **Facility Complexity 1a** | 93,817 (47.3%) | 74,743 (47.1%) | 19,074 (48.4%) | -0.026 |
| **VA Station Population** | 2,252.9 (1,181.95) | 2,254.0 (1,196.06) | 2,248.4 (1,123.33) | 0.005 |
| **Care Assessment Needs Score** | 51.6 (30.29) | 51.5 (30.30) | 52.1 (30.22) | -0.022 |
| Unknown | 2,883 | 2,253 | 630 |  |
| **Charlson Comorbidity Index** | 1.9 (2.27) | 1.9 (2.27) | 2.0 (2.30) | -0.020 |
| **Area Deprivation Index** | 53.2 (25.14) | 52.4 (25.38) | 56.5 (23.91) | -0.164 |
| Unknown | 519 | 429 | 90 |  |
| **Chronic Kidney Disease** | 32,615 (16.5%) | 26,101 (16.4%) | 6,514 (16.5%) | -0.002 |
| **Cardiovascular Disease** | 141,256 (71.3%) | 112,883 (71.1%) | 28,373 (72.0%) | -0.019 |
| **Diabetes** | 68,286 (34.5%) | 54,625 (34.4%) | 13,661 (34.6%) | -0.005 |
| **Immunocompromised** | 19,999 (10.1%) | 15,945 (10.0%) | 4,054 (10.3%) | -0.005 |
| **Liver Disease** | 19,851 (10.0%) | 15,647 (9.9%) | 4,204 (10.7%) | -0.027 |
| **Lung Disease** | 67,101 (33.9%) | 53,405 (33.6%) | 13,696 (34.7%) | -0.023 |
| **Substance/Alcohol Use Diagnosis** | 45,504 (23.0%) | 36,080 (22.7%) | 9,424 (23.9%) | -0.028 |
| **Mental Health Diagnosis** | 91,144 (46.0%) | 73,222 (46.1%) | 17,922 (45.5%) | 0.013 |
| **COVID-19 Vaccination Status** |  |  |  | 0.018 |
| Primary | 50,013 (25.2%) | 40,307 (25.4%) | 9,706 (24.6%) |  |
| Boosted | 106,943 (54.0%) | 85,485 (53.8%) | 21,458 (54.4%) |  |
| Unvaccinated | 41,218 (20.8%) | 32,956 (20.8%) | 8,262 (21.0%) |  |
| **30-day COVID-19 Hospitalization or Death** | 15,037 (7.6%) | 11,739 (7.4%) | 3,298 (8.4%) | -0.036 |
| **30-day All-cause Mortality** | 2,444 (1.2%) | 1,924 (1.2%) | 520 (1.3%) | -0.010 |
| **30-day COVID-19 Hospitalization** | 13,183 (6.7%) | 10,265 (6.5%) | 2,918 (7.4%) | -0.037 |

<sup>†</sup> Mean (SD); n (%)

### Model development

The ensemble models were successfully fitted with all ten pre-specified learners. In the composite and hospitalization models, only the XGBoost, random forest, and GAM learners received an ensemble weight; the XGBoost learner accounted for over 60% of the ensemble weights for the composite and hospitalization models. The mortality model showed greater diversity in learners that were weighted (including LASSO regression, and multivariate adaptive regression splines) with a more even weight distribution (Table 2).

**Table 2.** Ensemble model weights and 5-fold cross-validated area under the receiver operating characteristic curve (AUC) for individual learners and fitted ensemble model in training dataset.

| Ensemble Model Learner Weights |  |  |  | Mean 5-fold Cross-validated AUC (range) |  |  |
| --- | --- | --- | --- | --- | --- | --- |
|  | Composite model | Hospitalization model | Mortality model | Composite outcome model | Hospitalization model | Mortality model |
| SL Ensemble | -- | -- | -- | 0.821 (0.809, 0.834) | 0.803 (0.796, 0.811) | 0.911 (0.9, 0.925) |
| Mean Model | 0 | 0 | 0 | 0.5 (0.5, 0.5) | 0.5 (0.5, 0.5) | 0.5 (0.5, 0.5) |
| Logistic Regression | 0 | 0 | 0 | 0.813 (0.799, 0.825) | 0.796 (0.789, 0.801) | 0.904 (0.893, 0.916) |
| Bayesian GLM | 0 | <0.001 | 0 | 0.813 (0.800, 0.825) | 0.796 (0.789, 0.801) | 0.905 (0.893, 0.920) |
| MARS | 0 | 0 | 0.16 | 0.807 (0.794, 0.818) | 0.788 (0.782, 0.795) | 0.889 (0.873, 0.905) |
| GAM | 0.15 | 0.14 | 0.14 | 0.817 (0.805, 0.829) | 0.799 (0.792, 0.807) | 0.908 (0.899, 0.917) |
| Ridge Regression | 0 | 0 | 0 | 0.814 (0.800, 0.825) | 0.797 (0.79, 0.803) | 0.905 (0.893, 0.920) |
| LASSO Regression | 0 | 0 | 0.19 | 0.813 (0.800, 0.825) | 0.796 (0.789, 0.802) | 0.905 (0.893, 0.920) |
| Random Forest | 0.22 | 0.21 | 0.24 | 0.817 (0.803, 0.830) | 0.798 (0.791, 0.807) | 0.907 (0.893, 0.921) |
| Neural Network | 0 | 0 | 0 | 0.814 (0.801, 0.827) | 0.795 (0.787, 0.801) | 0.878 (0.854, 0.900) |
| XGBoost | 0.63 | 0.65 | 0.26 | 0.821 (0.809, 0.833) | 0.803 (0.796, 0.811) | 0.908 (0.897, 0.919) |

LM: generalized linear model; LASSO: least absolute shrinkage and selection operator.

Overall, each model showed good cross-validated discrimination with AUC estimates >0.80 in the composite (AUC: 0.82, range: 0.81 – 0.83) and hospitalization (AUC: 0.80, range: 0.80 – 0.81) models (Table 2). The mortality model had the highest cross-validated AUC of 0.91 (range: 0.90 – 0.93). The full ensemble models generally had higher average cross-validated AUCs than the individually included learners.

### Model performance in holdout test dataset

Histograms of predicted risks from each model are shown in Supplement S4, showing a right-skewed distribution with predicted risks ranging from <1% to 70% for the composite outcome model, <1% to 57% for the hospitalization model, and <1% to 49% for the mortality model. The ensemble models all performed well in the holdout test dataset with AUCs >0.80 and were similar to the cross-validated estimates (Figure 1). The Brier scores for the ensemble models were all close to 0 with the largest score in the composite model (Brier score: 0.07) and lowest score in the mortality model (Brier score: 0.01) suggesting good prediction and classification accuracy. The ensemble models all showed good model calibration, with calibration slopes all close to 1 suggesting the spread of estimated risks generally align with observed risk. Calibration intercepts were all positive suggesting slight underestimation in predicted risks, but the magnitude of miscalibration was low with calibration intercepts all <0.3 (Figure 1). Model calibration by age, sex, race, ethnicity, region, and CAN score generally display consistent calibration across all subgroups (see Supplement S5a – S5c).

**Figure 1.**
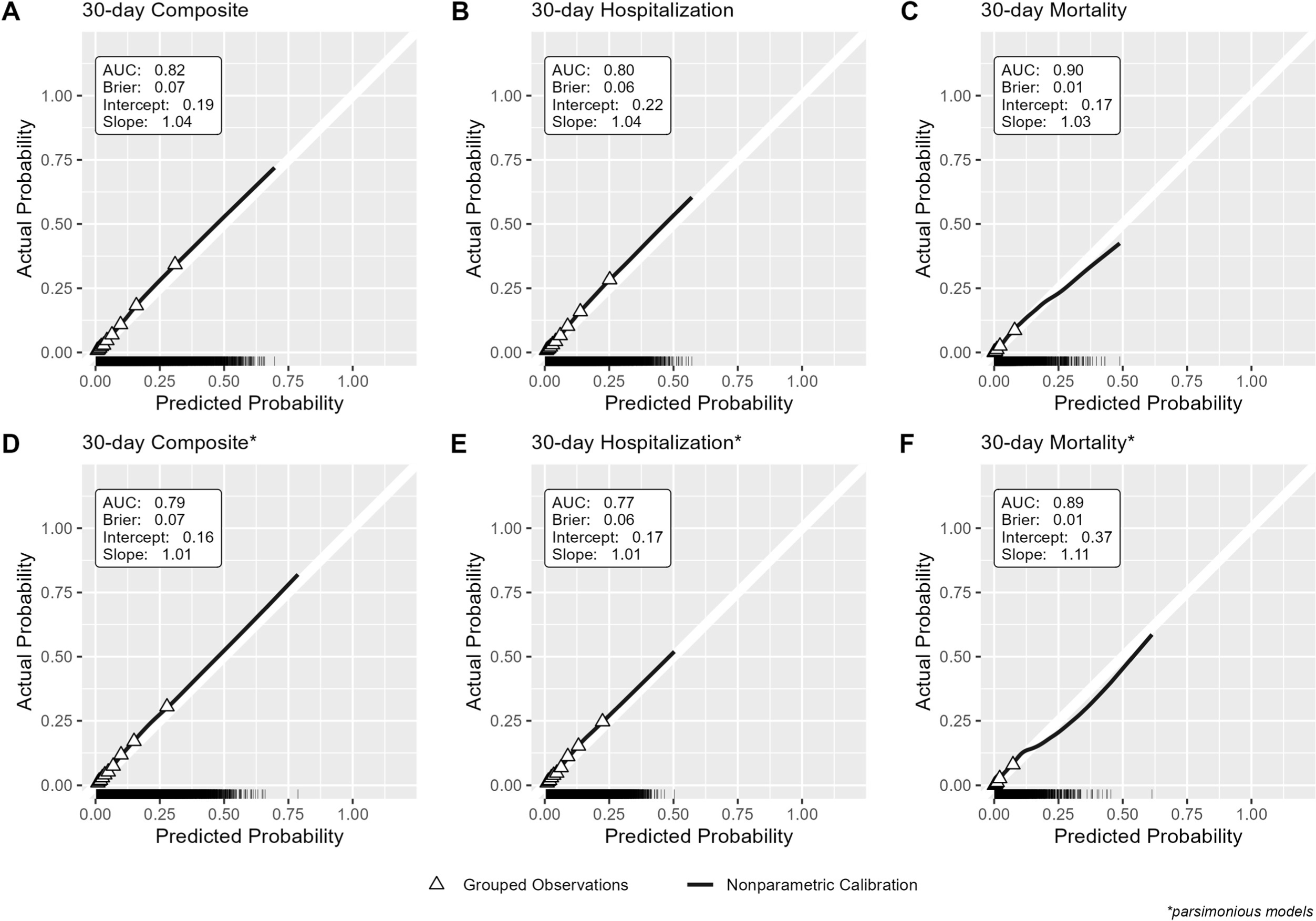
Calibration plots and performance metrics in holdout test dataset for A) Full 30-day Composite Model, B) Full 30-day Hospitalization Model, C) Full 30-day Mortality Model, D) Parsimonious 30-day Composite Model, E) Parsimonious 30-day Hospitalization Mode, and F) Parsimonious 30-day Mortality Model. White 45-degree line represents ideal model calibration. Solid black line represents the nonparametric local regression-smoothed calibration curve. Triangle points represent the average actual outcome probability for grouped deciles of the predicted probability. The rug plot at the bottom of each panel shows the distribution of predicted probabilities.

### Comparison with existing risk scores

Both the recalibrated CAN score and VACO Index had lower AUCs than our full ensemble models for predicting COVID-19 outcomes, although the AUCs for mortality in both the CAN score and VACO Index were >0.80 (Supplement S6). In terms of model calibration, the CAN score showed nearly perfect calibration with intercepts <0.2 and slopes of 1. The VACO Index had an intercept of −2.1 and a calibration slope >1, suggesting more extreme predicted risks than observed in our holdout cohort. Compared with the VACO Index, our mortality model showed an absolute increase of +13% in correctly reclassifying 30-day mortality (Supplement S7a). Compared with the recalibrated CAN score, our mortality model showed a decrease in the absolute NRI of −7%, driven by reclassification errors among patients without the outcome.

### Variable importance

SHAP values were estimated from the XGBoost model in each ensemble since it was the learner with the greatest weight and values for the top 15 variable domains in each ensemble model are shown in Figure 2 (See Supplement S7 for SHAP values for all predictors). In all models, the CAN score had the highest importance with SHAP values all >0.5 and was highest in the mortality model (CAN score SHAP value: 1.35). The vulnerability and frailty indices were strong predictors of outcomes with CAN, CCI, and ADI all included in the top 15 predictors. Outpatient COVID-19 treatment and vaccination status were important predictors, ranking among the top 5 with largest SHAP values. Facility complexity level was also among the top 5 predictors in the composite and hospitalization models (but not the mortality model), suggesting treating facility complexity may be more relevant for hospitalization risk than death (43). While cardiovascular disease, receipt of immunosuppressive medications or cancer therapies, and mental health conditions were among the top 15 predictors, other high-risk chronic conditions like diabetes, liver disease, and lung disease did not have high SHAP values (see Supplement for full list). Notably, race had the 9^th^ largest SHAP value (SHAP value: 0.076) in the mortality model but was not among the top 15 features for the composite or hospitalization models.

**Figure 2.**
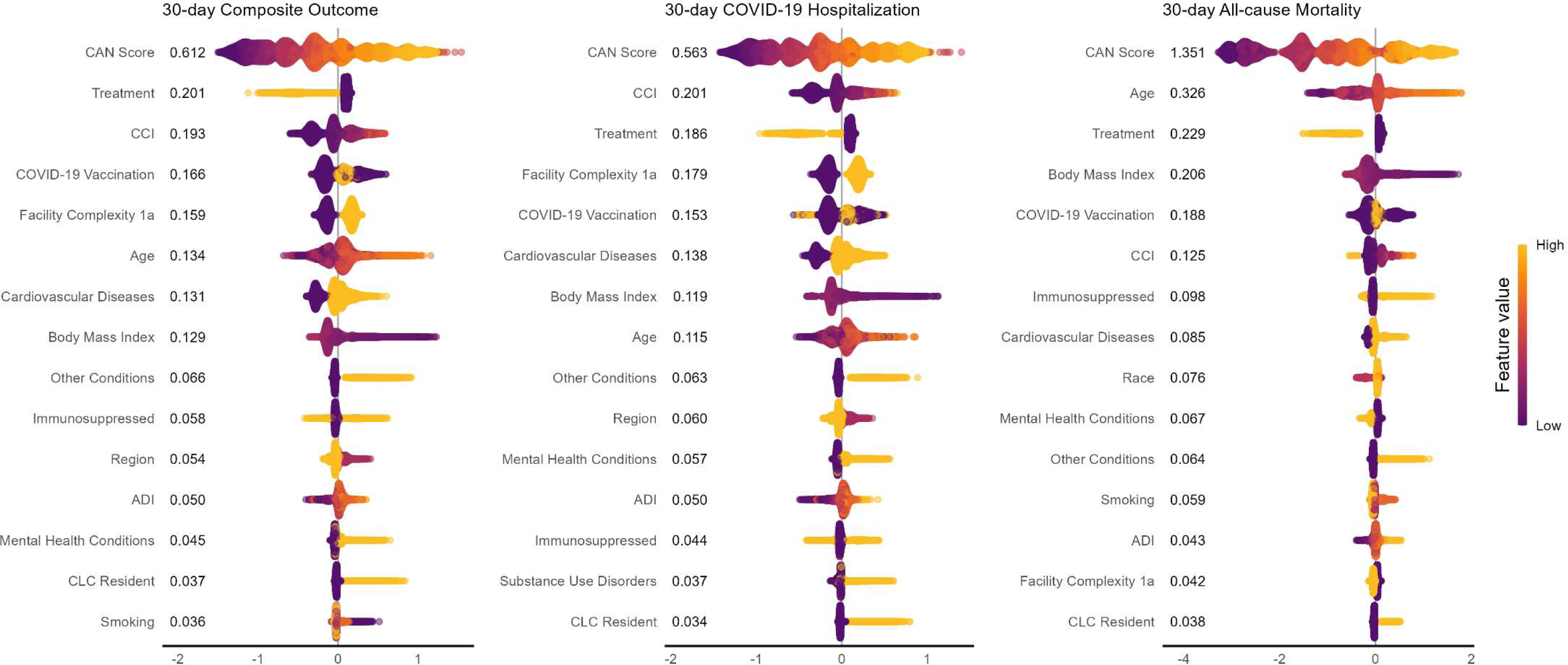
Bee swarm plot showing distribution of SHAP values and mean absolute SHAP value from outcome models predicted on holdout test dataset. ADI: area deprivation index CCI: Charlson Comorbidity Index CAN score: Care Assessment Needs score CLC: community living center Treatment SHAP value estimate includes indicators for receipt of outpatient nirmatrelvir-ritonavir, molnupiravir, or anti-SARS-CoV-2 monoclonal antibody treatment. Age includes age at index and indicator for being >65 years old. SHAP value estimates for cardiovascular diseases, immunocompromise, other diseases, substance use disorders, and mental health conditions, include SHAP values for all indicators within those domains (see supplement for list of indicators and their domains used in prediction).

### Parsimonious models

The performance and calibration of the parsimonious models trained using the subset of the top 15 most important predictors are shown in Supplement S9. Overall, these parsimonious models showed comparable discrimination to the fully fitted models (parsimonious composite model AUC: 0.79, parsimonious hospitalization model AUC: 0.77, and parsimonious mortality model AUC: 0.89) and slightly improved calibration in the composite and hospitalization model, but poorer calibration in the parsimonious model where risks were underestimated. The absolute NRIs for all parsimonious models compared with fully fitted models showed a loss in reclassification accuracy of about −2% in the composite and hospitalization model and a loss in reclassification accuracy of <1% in the mortality model (Supplement S11).

## DISCUSSION

Using data from a national cohort of veterans diagnosed with COVID-19 during the Omicron era, we developed three multivariable prediction models for 30-day COVID-19 hospitalization and death which demonstrated good discrimination and calibration in internal validation. We found age, composite measures of frailty including CCI and CAN score, select medical comorbidities such as cardiovascular disease, COVID-19 vaccination, and outpatient COVID-19 pharmacotherapies to be important predictors of adverse outcomes. Parsimonious models which included a small subset of predictors created to be accessible in non-VHA healthcare systems also showed comparable performance.

In all models, we found the CAN score to be the most important predictor for adverse COVID-19 outcomes. Osborne et al., previously demonstrated the CAN score was associated with COVID-19 mortality in the first three months of the pandemic and recommended its use for risk stratification (32). The CAN score is an automated and validated model for predicting patient hospitalization or death developed and used at the VHA (4). In addition to demographic and comorbidity data, the CAN score includes healthcare utilization measurements such emergency department visits and medication refills, and vital sign measurements like blood pressure and heart rate (31). While the CAN score was the most important predictor in our ensemble models and performed well alone once calibrated (Supplement S6), it is not available in healthcare systems outside of the VHA. Our parsimonious models, which excluded the CAN score, performed comparably to our fully fitted models and could be implemented more widely.

Our models have multiple strengths. They address a significant gap in the field, given that only 5% of developed COVID-19 outcome models predict hospitalization risk (3). We also focused on COVID-19 diagnoses during the Omicron era and included COVID-19 vaccines and treatments. Following the early pandemic and Delta era, COVID-19 hospitalization and mortality rates significantly declined, and effective vaccines and treatments became available. Thus, early risk prediction models may no longer be well-calibrated (44). Furthermore, many prior risk models were developed during a period of lower population immunity (45). Dickerman et al. have demonstrated how prediction model calibration can worsen when applied to settings where treatment strategies and availability differ from the context in which models were developed (14). We demonstrated that our prediction models outperformed the VACO Index (11), with improved discrimination, calibration, and classification for mortality. We chose not to assess the VACO Index’s performance on hospitalization outcomes since it was only developed and validated for mortality. Reasons for this improvement could be our use of an ensemble of model and a larger number of predictors; however, model training on a more recent cohort reflecting contemporary contexts may be the most important as we showed the VACO index is no longer well calibrated.

Our models have important implications for clinical practice and research applications. Since our models were trained using predictors readily available in CSDR, it is possible to automate risk scores for all VHA patients similar to the CAN score (4). Identifying high risk patients who might benefit the most from outpatient COVID-19 treatments, for example, could facilitate improved prescribing, particularly as utilization remains low in many settings (46,47). In addition, the parsimonious models, which exclude VHA-specific predictors, may be more broadly implemented in other healthcare systems as well as facilitate recruitment of high-risk patients to power future clinical trials.

### Limitations

Our risk models have several important limitations. First, we did not have data on re-infections, an important prognostic factor for severe COVID-19-related outcomes. Second, while we considered many predictors for adverse COVID-19 outcomes, it is possible we are missing important, unmeasured prognostic factors; however, such factors would also likely be unavailable for model implementation. Third, while we expect the models to perform well in an out-of-sample population given good cross-validated discrimination, we have not demonstrated external model validation. Since our models were developed using readily available predictors, validation in future infection periods is possible and external validation of our parsimonious models in a separate healthcare system is feasible. Finally, while we included a diverse set of learners in our ensemble models, our set of ten learners is relatively small compared with typical super learner applications (20). We limited the number of included learners for computational efficiency; future work could consider incorporating additional algorithms and models with multiple parameter sets to enhance prediction (22).

## CONCLUSION

We developed machine learning risk prediction models that accurately estimate risk of COVID-19 hospitalization and mortality in the VHA, in the context of recent, Omicron-era COVID-19 infections, COVID-19 vaccinations and antiviral treatments. These models may be used for risk stratification of individual patients to optimize care and to identify high-risk patients for inclusion in clinical trials.

## Data Availability

All data produced in the present study are available upon reasonable request to the authors

## Disclaimer

The contents of this article do not represent the views of the U.S. Department of Veterans Affairs or the U.S. government.

## Additional Contributions

We thank the Biomedical Advanced Research and Development Authority (interagency agreement No. AAI21050) and US Food and Drug Administration (interagency agreement No. 75F40121S30013) for their support. These institutions did not directly contribute to this study.

## REFERENCES

1. Wang L, Zhang Y, Wang D, Tong X, Liu T, Zhang S, et al. Artificial Intelligence for COVID-19: A Systematic Review. Front Med (Lausanne). 2021;8:704256.

2. Shakibfar S, Nyberg F, Li H, Zhao J, Nordeng HME, Sandve GKF, et al. Artificial intelligence-driven prediction of COVID-19-related hospitalization and death: a systematic review. Front Public Health. 2023;11:1183725.

3. Appel KS, Geisler R, Maier D, Miljukov O, Hopff SM, Vehreschild JJ. A Systematic Review of Predictor Composition, Outcomes, Risk of Bias, and Validation of COVID-19 Prognostic Scores. Clinical Infectious Diseases. 2023 Oct 25;ciad618.

4. Atkins D, Makridis CA, Alterovitz G, Ramoni R, Clancy C. Developing and Implementing Predictive Models in a Learning Healthcare System: Traditional and Artificial Intelligence Approaches in the Veterans Health Administration. Annu Rev Biomed Data Sci. 2022 Aug 10;5(1):393–413.

5. Ahmad FB, Cisewski JA, Xu J, Anderson RN. COVID-19 Mortality Update - United States, 2022. MMWR Morb Mortal Wkly Rep. 2023 May 5;72(18):493–6.

6. Bajema KL, Berry K, Streja E, Rajeevan N, Li Y, Mutalik P, et al. Effectiveness of COVID-19 Treatment With Nirmatrelvir–Ritonavir or Molnupiravir Among U.S. Veterans: Target Trial Emulation Studies With One-Month and Six-Month Outcomes. Ann Intern Med. 2023 Jun 20;176(6):807–16.

7. Hammond J, Leister-Tebbe H, Gardner A, Abreu P, Bao W, Wisemandle W, et al. Oral Nirmatrelvir for High-Risk, Nonhospitalized Adults with Covid-19. N Engl J Med. 2022 Apr 14;386(15):1397–408.

8. Barda N, Riesel D, Akriv A, Levy J, Finkel U, Yona G, et al. Developing a COVID-19 mortality risk prediction model when individual-level data are not available. Nat Commun. 2020 Sep 7;11(1):4439.

9. Cisterna-García A, Guillén-Teruel A, Caracena M, Pérez E, Jiménez F, Francisco-Verdú FJ, et al. A predictive model for hospitalization and survival to COVID-19 in a retrospective population-based study. Scientific Reports. 2022 Oct 28;12(1):18126.

10. Dashti H, Roche EC, Bates DW, Mora S, Demler O. SARS2 simplified scores to estimate risk of hospitalization and death among patients with COVID-19. Scientific Reports. 2021 Mar 2;11(1):4945.

11. King JT Jr, Yoon JS, Rentsch CT, Tate JP, Park LS, Kidwai-Khan F, et al. Development and validation of a 30-day mortality index based on pre-existing medical administrative data from 13,323 COVID-19 patients: The Veterans Health Administration COVID-19 (VACO) Index. PLOS ONE. 2020 Nov 11;15(11):e0241825.

12. Murtas R, Morici N, Cogliati C, Puoti M, Omazzi B, Bergamaschi W, et al. Algorithm for Individual Prediction of COVID-19–Related Hospitalization Based on Symptoms: Development and Implementation Study. JMIR Public Health Surveill. 2021 Nov 15;7(11):e29504.

13. Yuan Y, Sun C, Tang X, Cheng C, Mombaerts L, Wang M, et al. Development and Validation of a Prognostic Risk Score System for COVID-19 Inpatients: A Multi-Center Retrospective Study in China. Engineering (Beijing). 2022 Jan;8:116–21.

14. Dickerman BA, Dahabreh IJ, Cantos KV, Logan RW, Lodi S, Rentsch CT, et al. Predicting counterfactual risks under hypothetical treatment strategies: an application to HIV. European Journal of Epidemiology. 2022 Apr 1;37(4):367–76.

15. Subbaswamy A, Saria S. From development to deployment: dataset shift, causality, and shift-stable models in health AI. Biostatistics. 2020 Apr 1;21(2):345–52.

16. Gao Y, Cai GY, Fang W, Li HY, Wang SY, Chen L, et al. Machine learning based early warning system enables accurate mortality risk prediction for COVID-19. Nature Communications. 2020 Oct 6;11(1):5033.

17. Horvath A, Lind T, Frece N, Wurzer H, Stadlbauer V. Validation of a simple risk stratification tool for COVID-19 mortality. Frontiers in Medicine [Internet]. 2022;9. Available from: https://www.frontiersin.org/articles/10.3389/fmed.2022.1016180

18. Knight SR, Ho A, Pius R, Buchan I, Carson G, Drake TM, et al. Risk stratification of patients admitted to hospital with covid-19 using the ISARIC WHO Clinical Characterisation Protocol: development and validation of the 4C Mortality Score. BMJ. 2020 Sep 9;370:m3339.

19. Goodacre S, Thomas B, Sutton L, Burnsall M, Lee E, Bradburn M, et al. Derivation and validation of a clinical severity score for acutely ill adults with suspected COVID-19: The PRIEST observational cohort study. PLOS ONE. 2021 Jan 22;16(1):e0245840.

20. van der Laan MJ, Polley EC, Hubbard AE. Super learner. Stat Appl Genet Mol Biol. 2007;6:Article25.

21. Naimi AI, Balzer LB. Stacked generalization: an introduction to super learning. European Journal of Epidemiology. 2018 May 1;33(5):459–64.

22. Phillips RV, van der Laan MJ, Lee H, Gruber S. Practical considerations for specifying a super learner. International Journal of Epidemiology. 2023 Aug 1;52(4):1276–85.

23. Collins GS, Reitsma JB, Altman DG, Moons KGM. Transparent Reporting of a multivariable prediction model for Individual Prognosis Or Diagnosis (TRIPOD): The TRIPOD Statement. Ann Intern Med. 2015 Jan 6;162(1):55–63.

24. Admon AJ, Wander PL, Iwashyna TJ, Ioannou GN, Boyko EJ, Hynes DM, et al. Consensus elements for observational research on COVID-19-related long-term outcomes. Medicine. 2022;101(46):8.

25. Ioannou GN, Green P, Fan VS, Dominitz JA, O’Hare AM, Backus LI, et al. Development of COVIDVax Model to Estimate the Risk of SARS-CoV-2–Related Death Among 7.6 Million US Veterans for Use in Vaccination Prioritization. JAMA Network Open. 2021 Apr 6;4(4):e214347–e214347.

26. Ioannou GN, O’Hare AM, Berry K, Fan VS, Crothers K, Eastment MC, et al. Trends Over Time in the Risk of Adverse Outcomes Among Patients With Severe Acute Respiratory Syndrome Coronavirus 2 Infection. Clin Infect Dis. 2022 Feb 11;74(3):416–26.

27. Pajouheshnia R, Peelen LM, Moons KGM, Reitsma JB, Groenwold RHH. Accounting for treatment use when validating a prognostic model: a simulation study. BMC Medical Research Methodology. 2017 Jul 14;17(1):103.

28. Groenwold RHH, Moons KGM, Pajouheshnia R, Altman DG, Collins GS, Debray TPA, et al. Explicit inclusion of treatment in prognostic modeling was recommended in observational and randomized settings. Journal of Clinical Epidemiology. 2016 Oct 1;78:90–100.

29. Ghirimoldi FM, Schmidt S, Simon RC, Wang CP, Wang Z, Brimhall BB, et al. Association of Socioeconomic Area Deprivation Index with Hospital Readmissions After Colon and Rectal Surgery. J Gastrointest Surg. 2021 Mar;25(3):795–808.

30. Charlson ME, Pompei P, Ales KL, MacKenzie CR. A new method of classifying prognostic comorbidity in longitudinal studies: Development and validation. Journal of Chronic Diseases. 1987 Jan 1;40(5):373–83.

31. Wang L, Porter B, Maynard C, Evans G, Bryson C, Sun H, et al. Predicting Risk of Hospitalization or Death Among Patients Receiving Primary Care in the Veterans Health Administration. Medical Care [Internet]. 2013;51(4). Available from: https://journals.lww.com/lww-medicalcare/fulltext/2013/04000/predicting_risk_of_hospitalization_or_death_among.12.aspx

32. Osborne TF, Veigulis ZP, Arreola DM, Röösli E, Curtin CM. Automated EHR score to predict COVID-19 outcomes at US Department of Veterans Affairs. PLOS ONE. 2020 Jul 27;15(7):e0236554.

33. Gruber S, Lee H, Phillips R, Ho M, van der Laan M. Developing a Targeted Learning-Based Statistical Analysis Plan. Statistics in Biopharmaceutical Research. 2023 Jul 3;15(3):468– 75.

34. Zachary Butzin-Dozier, Yunwen Ji, Haodong Li, Jeremy Coyle, Junming (Seraphina) Shi, Rachael V. Philips, et al. Predicting Long COVID in the National COVID Cohort Collaborative Using Super Learner. medRxiv. 2023 Jan 1;2023.07.27.23293272.

35. Pfaff ER, Girvin AT, Bennett TD, Bhatia A, Brooks IM, Deer RR, et al. Identifying who has long COVID in the USA: a machine learning approach using N3C data. The Lancet Digital Health. 2022 Jul 1;4(7):e532–41.

36. LeDell E, van der Laan MJ, Petersen M. AUC-Maximizing Ensembles through Metalearning. 2016;12(1):203–18.

37. Kuhn M. Building Predictive Models in R Using the caret Package. J Stat Soft. 2008 Nov 10;28(5):1–26.

38. Austin PC, Steyerberg EW. Graphical assessment of internal and external calibration of logistic regression models by using loess smoothers. Stat Med. 2014 Feb 10;33(3):517–35.

39. Huang Y, Li W, Macheret F, Gabriel RA, Ohno-Machado L. A tutorial on calibration measurements and calibration models for clinical prediction models. Journal of the American Medical Informatics Association. 2020 Apr 1;27(4):621–33.

40. MDCalc [Internet]. [cited 2023 Nov 3]. Veterans Health Administration COVID-19 (VACO) Index for COVID-19 Mortality. Available from: https://www.mdcalc.com/calc/10346/veterans-health-administration-covid-19-vaco-index-covid-19-mortality

41. Alba AC, Agoritsas T, Walsh M, Hanna S, Iorio A, Devereaux PJ, et al. Discrimination and Calibration of Clinical Prediction Models: Users’ Guides to the Medical Literature. JAMA. 2017 Oct 10;318(14):1377–84.

42. Lundberg SM, Lee SI. A unified approach to interpreting model predictions. Advances in neural information processing systems. 2017;30.

43. National Academies of Sciences E, Education D of B and SS and, Integration B on HS, Sciences D on E and P, Environment B on I and the C, Administration C on FSR for VH. Nature of Veterans Health Administration Facilities Management (Engineering) Tasks and Staffing. In: Facilities Staffing Requirements for the Veterans Health Administration-Resource Planning and Methodology for the Future [Internet]. National Academies Press (US); 2019 [cited 2023 Nov 6]. Available from: https://www.ncbi.nlm.nih.gov/books/NBK555777/

44. Finlayson SG, Subbaswamy A, Singh K, Bowers J, Kupke A, Zittrain J, et al. The Clinician and Dataset Shift in Artificial Intelligence. N Engl J Med. 2021 Jul 15;385(3):283–6.

45. CDC. Centers for Disease Control and Prevention. 2020 [cited 2023 Nov 1]. COVID Data Tracker. Available from: https://covid.cdc.gov/covid-data-tracker/index.html#nationwide-blood-donor-seroprevalence

46. Yan L, Streja E, Li Y, Rajeevan N, Rowneki M, Berry K, et al. Anti–SARS-CoV-2 Pharmacotherapies Among Nonhospitalized US Veterans, January 2022 to January 2023. JAMA Netw Open. 2023 Aug 31;6(8):e2331249.

47. Sullivan M, Perrine CG, Kelleher J, Kanwar O, Kuwabara S, Bennett K, et al. Notes From the Field: Dispensing of Oral Antiviral Drugs for Treatment of COVID-19 by Zip Code-Level Social Vulnerability - United States, December 23, 2021-August 28, 2022. MMWR Morb Mortal Wkly Rep. 2022 Oct 28;71(43):1384–5.

