## Supplement for "Development of a prediction model for 30-day COVID-19 hospitalization and death in a national cohort of Veterans Health Administration patients – March 2022 - April 2023"

Supplemental Tables and Figures

### Supplemental Tables and Figures

#### S1. List of processed predictors used for in all outcome models.

Note, 84 variables were included for modeling. After data pre-processing (one-hot coding categorical variables and creating missing/unknown indicators for missing values) we included a total of 99 predictors shown here in each model. All comorbidity indicators were for diagnoses within 2 years prior to infection unless otherwise specified.

| No. | Domain | Predictor | Type |
|---|---|---|---|
| 1 | Conditions/Risk Factors | Body Mass Index (kg/m2) | Continuous |
| 2 | Conditions/Risk Factors | Body Mass Index (kg/m2) - Unknown | Binary (0/1) |
| 3 | Comorbidity/Vulnerability Scores | Charlson Comorbidity Index (CCI) | Continuous |
| 4 | Comorbidity/Vulnerability Scores | CAN Score (1-year mortality risk) | Continuous |
| 5 | Comorbidity/Vulnerability Scores | CAN Score (1-year mortality risk) - Unknown | Binary (0/1) |
| 6 | Comorbidity/Vulnerability Scores | Area Deprivation Index (ADI) | Continuous |
| 7 | Comorbidity/Vulnerability Scores | Area Deprivation Index (ADI) - Unknown | Binary (0/1) |
| 8 | Conditions/Risk Factors | Stroke/Cerebrovascular - Hemorrhagic Stroke | Binary (0/1) |
| 9 | Conditions/Risk Factors | Stroke/Cerebrovascular - Ischemic Stroke | Binary (0/1) |
| 10 | Conditions/Risk Factors | Stroke/Cerebrovascular - Cerebrovascular disease | Binary (0/1) |
| 11 | Conditions/Risk Factors | Stroke/Cerebrovascular - any (composite) | Binary (0/1) |
| 12 | Conditions/Risk Factors | Liver disease - Chronic Hepatitis | Binary (0/1) |
| 13 | Conditions/Risk Factors | Liver disease - Cirrhosis | Binary (0/1) |
| 14 | Conditions/Risk Factors | Liver disease - Liver disease | Binary (0/1) |
| 15 | Conditions/Risk Factors | Liver disease - Mild liver disease | Binary (0/1) |
| 16 | Conditions/Risk Factors | Liver disease - Moderate-to-severe liver disease | Binary (0/1) |
| 17 | Conditions/Risk Factors | Liver disease - any (composite) | Binary (0/1) |
| 18 | Conditions/Risk Factors | Lung disease - Asthma | Binary (0/1) |
| 19 | Conditions/Risk Factors | Lung disease - Chronic lung disease | Binary (0/1) |
| 20 | Conditions/Risk Factors | Lung disease - COPD | Binary (0/1) |
| 21 | Conditions/Risk Factors | Lung disease - Emphysema | Binary (0/1) |
| 22 | Conditions/Risk Factors | Lung disease - Pulmonary Fibrosis | Binary (0/1) |
| 23 | Conditions/Risk Factors | Lung disease - PulmHD2 | Binary (0/1) |
| 24 | Conditions/Risk Factors | Lung disease - any (composite) | Binary (0/1) |
| 25 | Conditions/Risk Factors | Smoking status - Current | Binary (0/1) |
| 26 | Conditions/Risk Factors | Smoking status - Former | Binary (0/1) |
| 27 | Conditions/Risk Factors | Smoking status - Unknown | Binary (0/1) |
| 28 | Conditions/Risk Factors | Cardiovascular disease - Acute MI | Binary (0/1) |
| 29 | Conditions/Risk Factors | Cardiovascular disease - Coronary Atherosclerosis/other Heart Disease | Binary (0/1) |
| 30 | Conditions/Risk Factors | Cardiovascular disease - Cardiomyopathy | Binary (0/1) |
| 31 | Conditions/Risk Factors | Cardiovascular disease - Congestive Heart Failure | Binary (0/1) |
| 32 | Conditions/Risk Factors | Cardiovascular disease - Chronic Rheumatic Heart Disease | Binary (0/1) |
| 33 | Conditions/Risk Factors | Cardiovascular disease - Cardiovascular Disease | Binary (0/1) |
| 34 | Conditions/Risk Factors | Cardiovascular disease - Heart Disease | Binary (0/1) |
| 35 | Conditions/Risk Factors | Cardiovascular disease - Heart Failure | Binary (0/1) |
| 36 | Conditions/Risk Factors | Cardiovascular disease - Hypertension | Binary (0/1) |
| 37 | Conditions/Risk Factors | Cardiovascular disease - Other heart disease | Binary (0/1) |
| 38 | Conditions/Risk Factors | Cardiovascular disease - Peripheral arterial disease | Binary (0/1) |
| 39 | Conditions/Risk Factors | Cardiovascular disease - any (composite) | Binary (0/1) |
| 40 | Conditions/Risk Factors | Kidney Disease - Chronic Kidney disease | Binary (0/1) |
| 41 | Conditions/Risk Factors | Kidney Disease - Chronic Kidney Failure | Binary (0/1) |
| 42 | Conditions/Risk Factors | Kidney Disease - Dialysis in past 2 years | Binary (0/1) |
| 43 | Conditions/Risk Factors | Kidney Disease - Dialysis at Index | Binary (0/1) |
| 44 | Conditions/Risk Factors | Kidney Disease - Nephrosis | Binary (0/1) |
| 45 | Conditions/Risk Factors | Kidney Disease - any (composite) | Binary (0/1) |
| 46 | Conditions/Risk Factors | Immunosuppressive condition - HIV | Binary (0/1) |
| 47 | Conditions/Risk Factors | Immunosuppressive condition - Cancer | Binary (0/1) |
| 48 | Conditions/Risk Factors | Immunosuppressive condition - Metastatic tumor | Binary (0/1) |
| 49 | Conditions/Risk Factors | Immunosuppressive condition - other (composite) | Binary (0/1) |
| 50 | Conditions/Risk Factors | Immunosuppressive condition - any (composite) | Binary (0/1) |
| 51 | Conditions/Risk Factors | Immunosuppressive medications | Binary (0/1) |

| | | | |
|---|---|---|---|
| 52 | Conditions/Risk Factors | Other type of Diabetes | Binary (0/1) |
| 53 | Conditions/Risk Factors | Diabetes - Type 1 | Binary (0/1) |
| 54 | Conditions/Risk Factors | Diabetes - Type 2 | Binary (0/1) |
| 55 | Conditions/Risk Factors | Diabetes with complication | Binary (0/1) |
| 56 | Conditions/Risk Factors | Diabetes without complication | Binary (0/1) |
| 57 | Conditions/Risk Factors | Diabetes - any (composite) | Binary (0/1) |
| 58 | Conditions/Risk Factors | Mental health conditions - Bipolar | Binary (0/1) |
| 59 | Conditions/Risk Factors | Mental health conditions - Depression | Binary (0/1) |
| 60 | Conditions/Risk Factors | Mental health conditions - Schizophrenia | Binary (0/1) |
| 61 | Conditions/Risk Factors | Mental health conditions - PTSD | Binary (0/1) |
| 62 | Conditions/Risk Factors | Mental health conditions - any mental health (composite) | Binary (0/1) |
| 63 | Conditions/Risk Factors | Substance/Alcohol Use - substance use disorder diagnosis | Binary (0/1) |
| 64 | Conditions/Risk Factors | Substance/Alcohol Use - alcohol use disorder diagnosis | Binary (0/1) |
| 65 | Conditions/Risk Factors | Substance/Alcohol Use - any substance use (composite) | Binary (0/1) |
| 66 | Conditions/Risk Factors | Other conditions - Chronic Neuromuscular Disease | Binary (0/1) |
| 67 | Conditions/Risk Factors | Other conditions - Epilepsy | Binary (0/1) |
| 68 | Conditions/Risk Factors | Other conditions - multiple sclerosis | Binary (0/1) |
| 69 | Conditions/Risk Factors | Other conditions - Parkinson’s | Binary (0/1) |
| 70 | Conditions/Risk Factors | Other conditions - Blood transfusion | Binary (0/1) |
| 71 | Conditions/Risk Factors | Other conditions - any (composite) | Binary (0/1) |
| 72 | Conditions/Risk Factors | Age ≥65 | Binary (0/1) |
| 73 | Conditions/Risk Factors | Overweight (body mass index ≥ 25) | Binary (0/1) |
| 74 | Conditions/Risk Factors | Pregnant at infection | Binary (0/1) |
| 75 | Conditions/Risk Factors | Blood - Sickle Cell | Binary (0/1) |
| 76 | Conditions/Risk Factors | Blood - Thalassemia | Binary (0/1) |
| 77 | Geography | Region - West | Binary (0/1) |
| 78 | Geography | Region - Midwest | Binary (0/1) |
| 79 | Geography | Region - South | Binary (0/1) |
| 80 | Geography | Urban residence | Binary (0/1) |
| 81 | Geography | Urban residence - Unknown | Binary (0/1) |
| 82 | Geography | VHA Index Facility Complexity 1a | Binary (0/1) |
| 83 | Demographic | Community Living Center Resident | Binary (0/1) |
| 84 | Demographic | Race - American Indian Alaska Native | Binary (0/1) |
| 85 | Demographic | Race - Asian | Binary (0/1) |
| 86 | Demographic | Race - Black | Binary (0/1) |
| 87 | Demographic | Race - Native Hawaiian Pacific Islander | Binary (0/1) |
| 88 | Demographic | Race - Unknown | Binary (0/1) |
| 89 | Demographic | Ethnicity - Hispanic or Latino | Binary (0/1) |
| 90 | Demographic | Ethnicity - Unknown | Binary (0/1) |
| 91 | Demographic | Age at index (years) | Continuous |
| 92 | Demographic | Age at index (years) - Unknown | Binary (0/1) |
| 93 | Demographic | Male Sex | Binary (0/1) |
| 94 | Vaccine/Treatment | Treatment - Receipt of any monoclonal antibodies | Binary (0/1) |
| 95 | Vaccine/Treatment | Treatment - Receipt of nirmatrelvir-ritonavir | Binary (0/1) |
| 96 | Vaccine/Treatment | Treatment - Receipt of molnupiravir | Binary (0/1) |
| 97 | Vaccine/Treatment | COVID-19 Vaccination - Primary Series | Binary (0/1) |
| 98 | Vaccine/Treatment | COVID-19 Vaccination - Boosted | Binary (0/1) |
| 99 | Vaccine/Treatment | Days since last vaccination | Continuous |

#### S2. Super Learner ensemble library and hyperparameter specifications

| Learner | R package (version) | Parameters/notes |
|---|---|---|
| Mean outcome | N/A | Classification on outcome average; included for benchmarking. |
| Generalized linear model | stats (4.3.1) | Main effects model; logistic regression |
| LASSO regression | glmnet (4.1-8) | Main effects model; default settings (alpha=1) |
| Ridge regression | glmnet (4.1-8) | Main effects model; default settings (alpha=0) |
| Bayesian GLM | arm (1.13-1) | Bayesian generalized linear model (logistic) with default settings |
| Generalized additive model | gam (1.22-2) | degree = 12 |
| Extreme gradient boosting | xgboost (1.7.5.1) | eta = 0.05, max_depth = 8, gamma=10, ntrees = 1200, minobspernode = 1 |
| Random forest | ranger (0.15.1) | num.trees = 1000, mtry = 20, min.node.size = 90, splitrule = "hellinger" |
| Neural network | nnet (7.3-19) | Size = 6, decay = 5, maxit = 500, MaxNWts = 3000 |
| Multivariate Adaptive Regression Splines | earth (5.3.2) | degree = 1, penalty = 3, nprune=18 |

#### S3. Super Learner ensemble specifications for each outcome model

| Model specifications | Composite outcome model | Hospitalization only model | Mortality only model |
|---|---|---|---|
| Dependent variable ($Y$) | 30-day COVID-19 hospitalization or death (binary outcome) | 30-day COVID-19 hospitalization (binary outcome) | 30-day all-cause mortality (binary outcome) |
| Predictors ($X$) | 84 categorical, binary, and continuous predictors* | Same | Same |
| Prediction function | $P(Y = 1 \mid X)$ | Same | Same |
| Performance metric for discrete Super Learner | Area Under the ROC Curve (AUC) | Same | Same |
| Meta learner | Non-negative binomial likelihood maximization | Same | Same |
| Training dataset sample size, $n$ | $n$ = 158,748 | Same | Same |
| Outcome prevalence ($\overline{p}$) | $\overline{p} = 0.0739$ | $\overline{p} = 0.0647$ | $\overline{p} = 0.0121$ |
| Effective sample size, $n_{eff}$** | $n_{eff} = 58{,}695$ | $n_{eff} = 51{,}325$ | $n_{eff} = 9{,}620$ |
| Minimum V-fold cross-validation (CV) recommended† | V ≥ 2 | V ≥ 2 | V ≥ 5 |
| V-fold cross-validation (CV) scheme implemented | Stratified V-fold CV (stratified on $Y$) with 5 folds | Same | Same |
| Learners considered in the SL library†† | Mean, GLM, Bayesian GLM, GAM, LASSO regression, ridge regression, random forest, neural network, and MARS. | Same | Same |

* See supplement for list and specification of predictors

** $n_{eff} = min(n, 5 * n_{rare});\ n_{rare} = n * min(\overline{p}, 1 - \overline{p})$

† Based on Phillips et al., 2023; $V \geq 2\ if\ n_{eff} \geq 10{,}000\ and\ V \geq 5\ if\ n_{eff} \geq 5{,}000\ and\ n_{eff} < 10{,}000$

†† GLM = generalized linear regression (logistic regression), GAM = generalized additive model, LASSO = least absolute shrinkage and selection operator, MARS = multivariate adaptive regression splines.

Phillips RV, van der Laan MJ, Lee H, Gruber S. Practical considerations for specifying a super learner. International Journal of Epidemiology. 2023 Aug 1;52(4):1276–85.

S4. Histogram showing distribution of predicted risks for A) 30-day composite outcome, B) 30-day hospitalization, and C) 30-day mortality in holdout sample color coded by empirical quartiles.

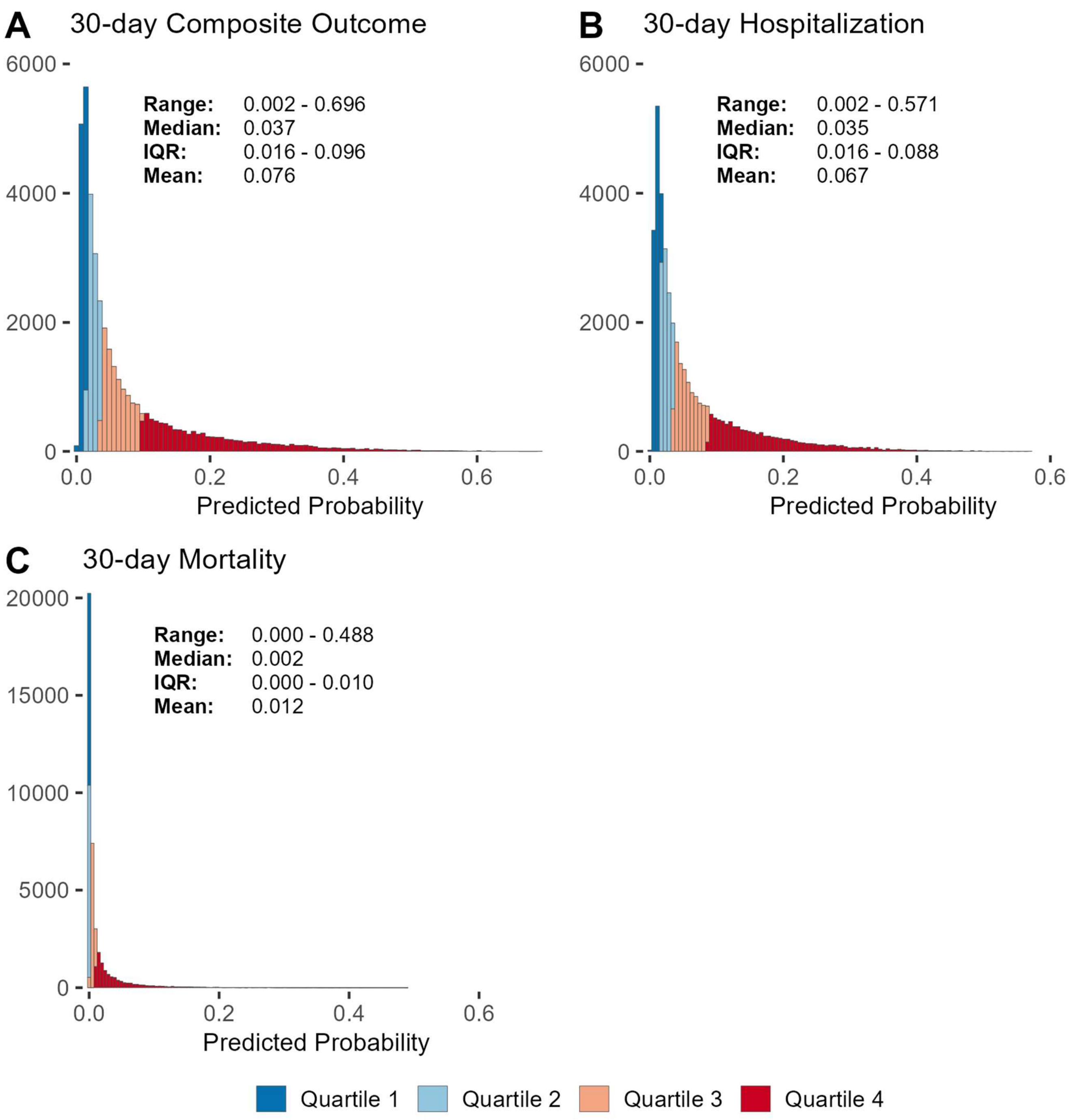

#### S5a. Calibration plots by sex, age, ethnicity, race, region, and CAN score - composite model

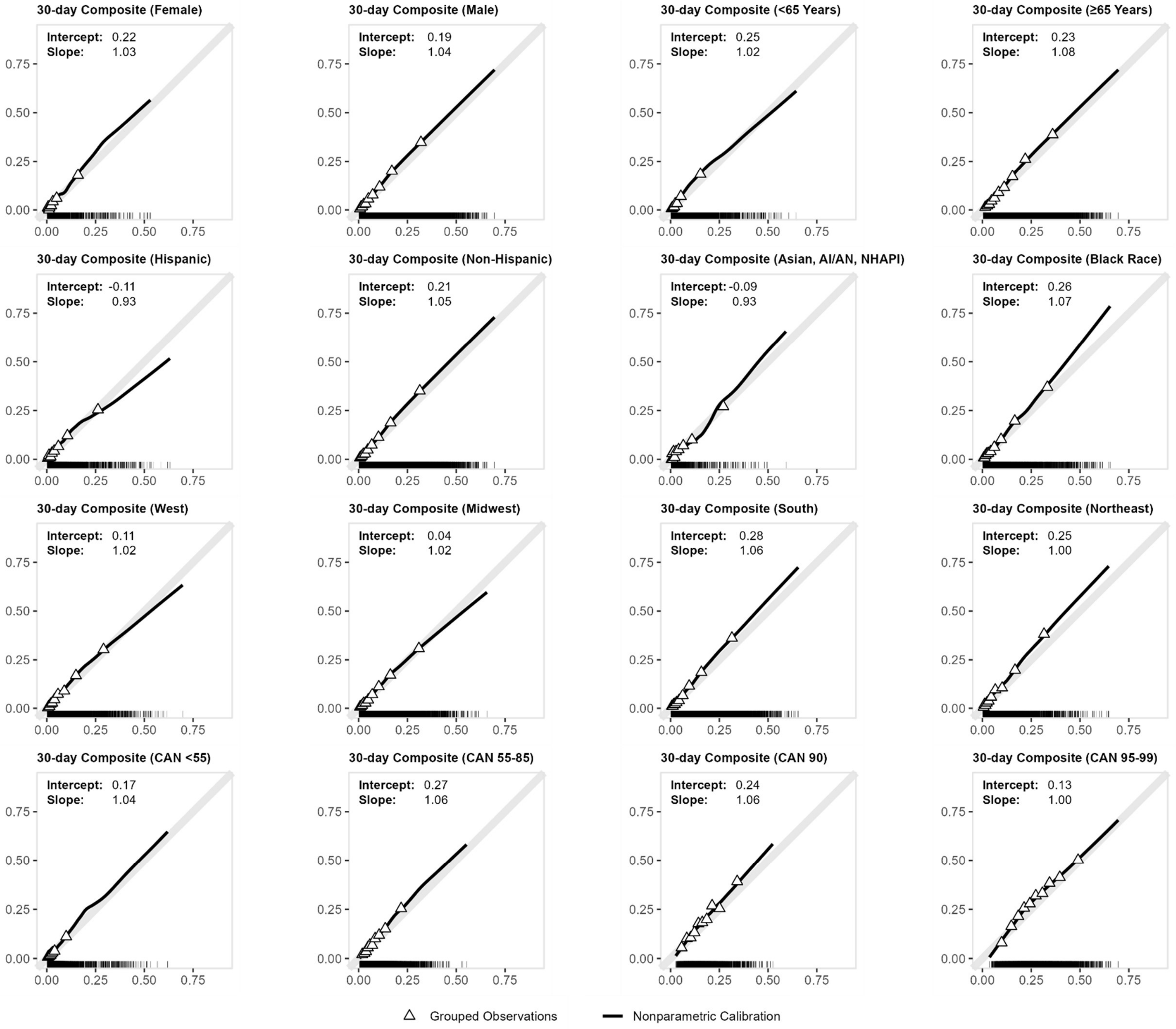

S5b. Calibration plots by sex, age, ethnicity, race, region, and CAN score subgroup - hospitalization model.

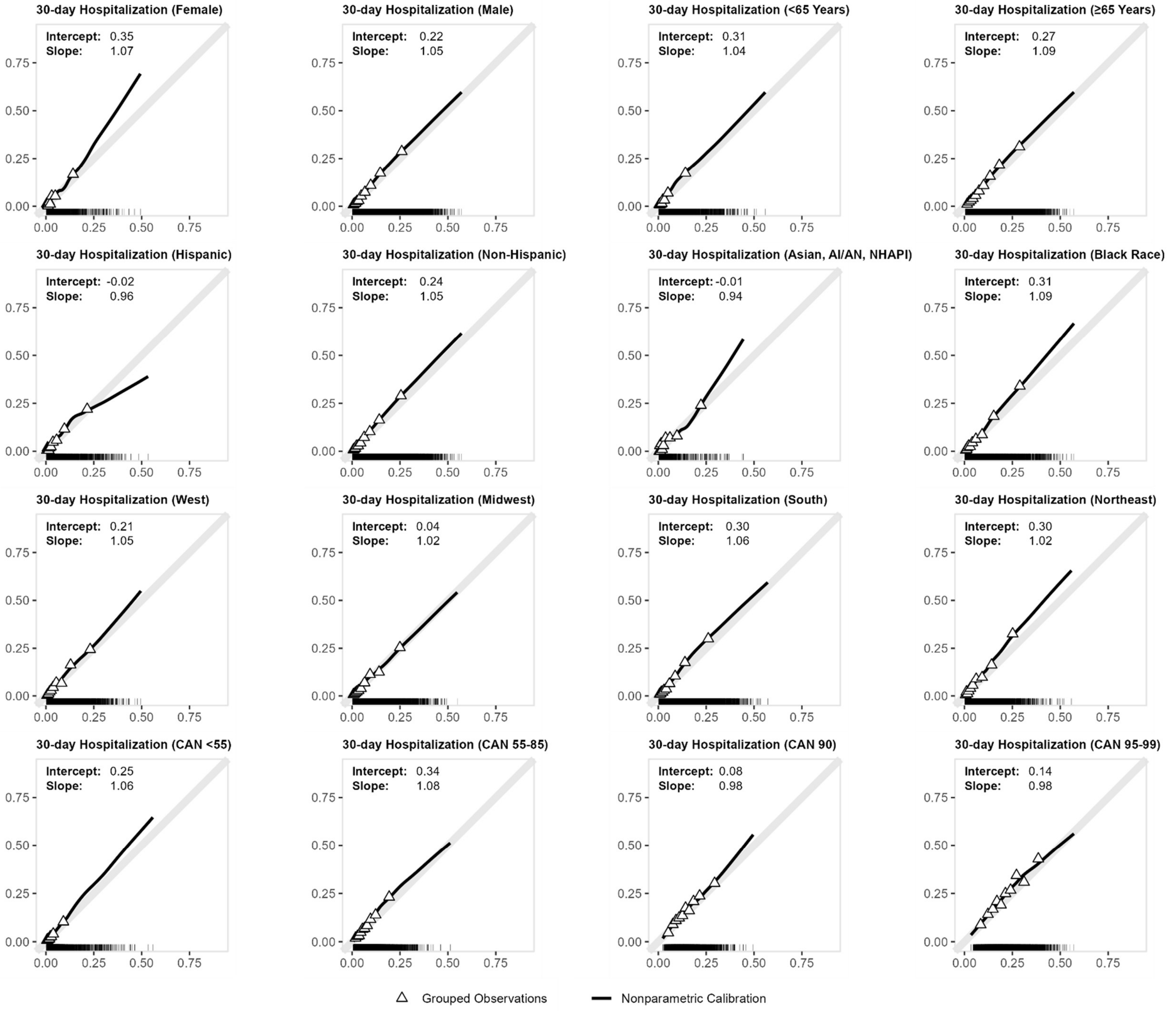

S5C. Calibration plots by sex, age, ethnicity, race, region, and CAN score subgroup - mortality model

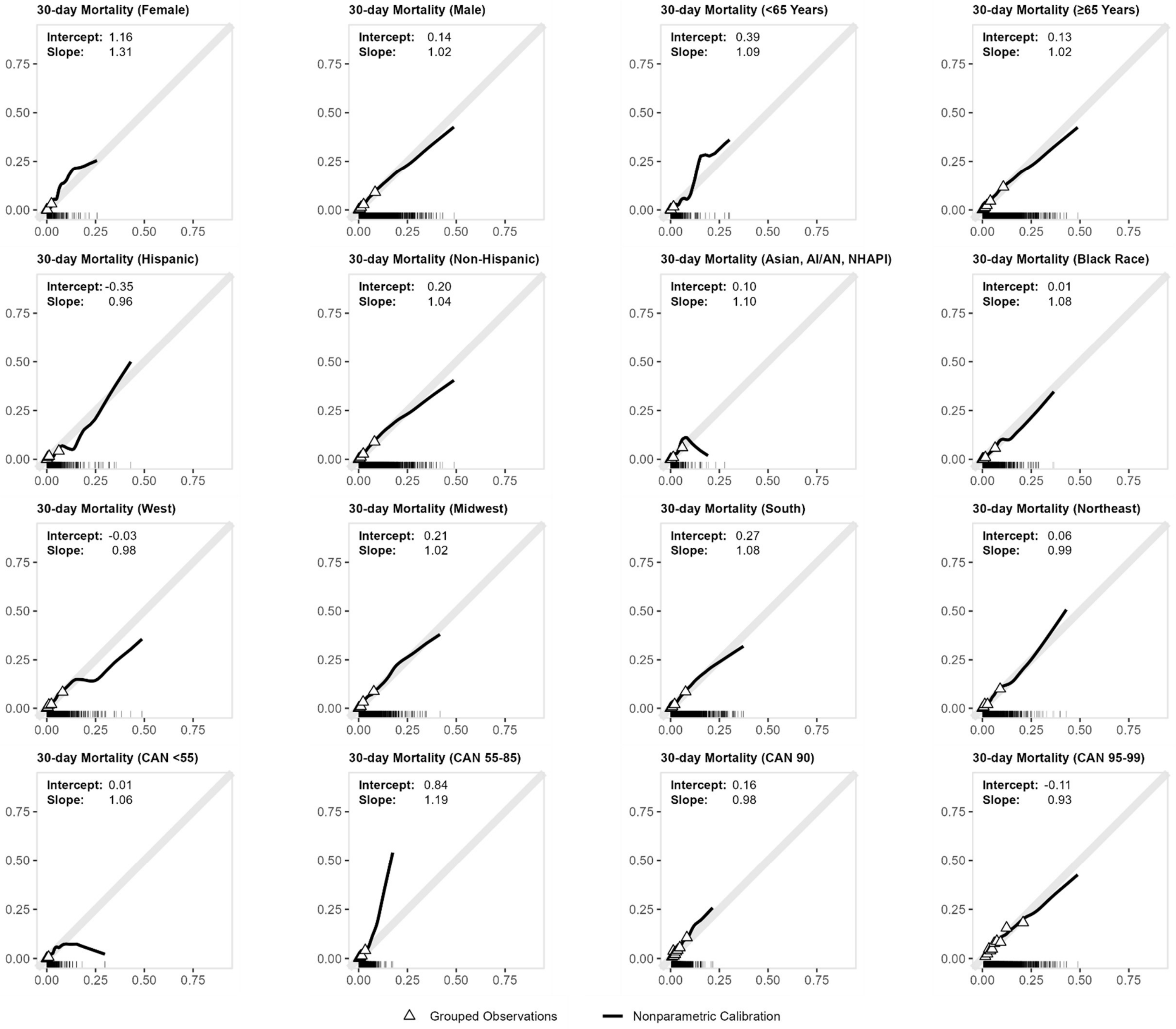

#### S6. The calibration plots of existing risk stratification models including the A-C) Care Assessment Need Score (CAN) and D) VACO index.

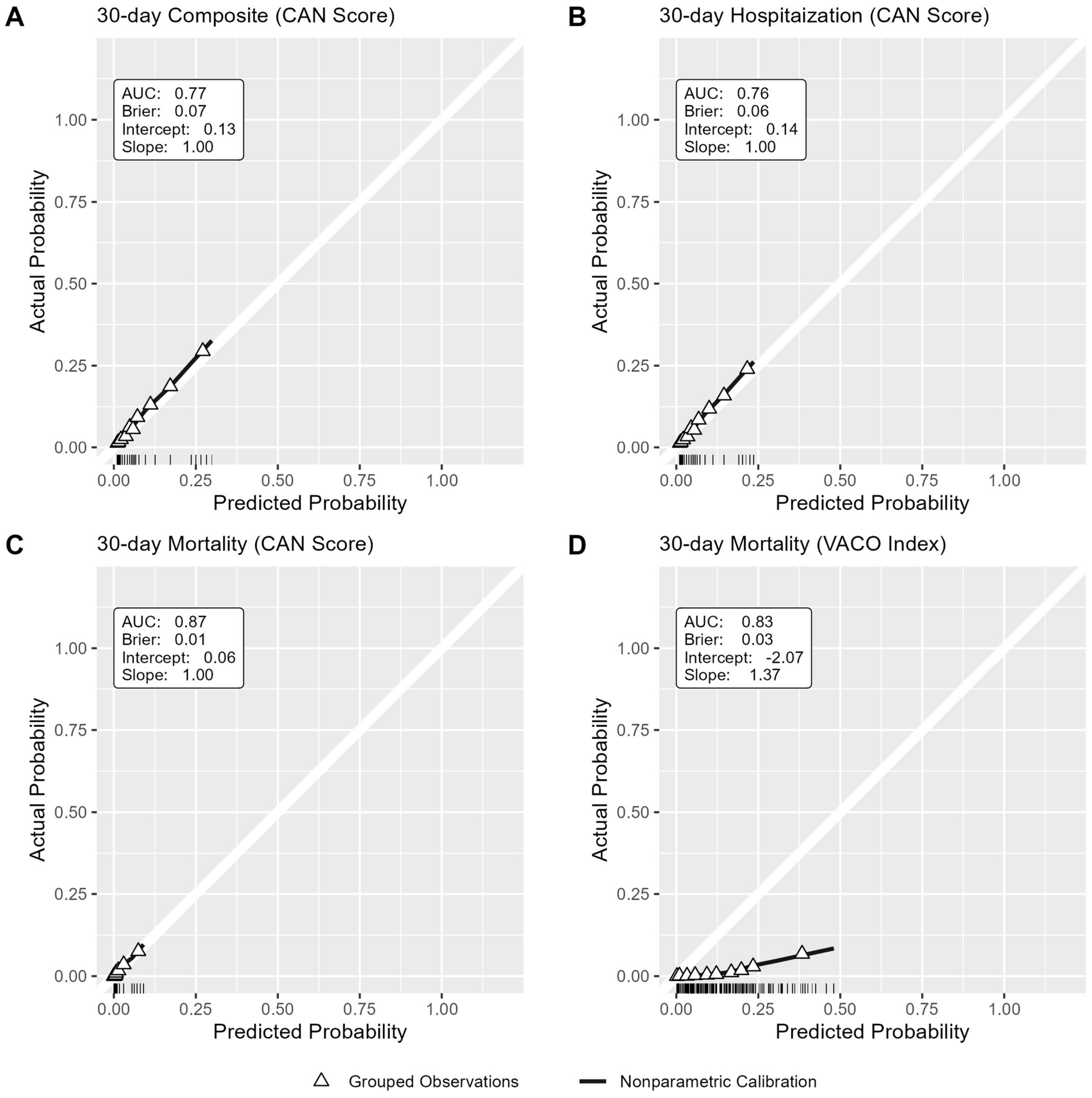

White 45-degree line represents ideal model calibration. Solid black line represents the nonparametric local regression-smoothed calibration curve. Triangle points represent the average actual outcome probability for grouped deciles of the predicted probability. The rug plot at the bottom of each panel shows the distribution of predicted probabilities.

#### S7a. Net Reclassification Index (NRI) of the 30-day all-cause mortality compared with the VACO Index

| 30-day Mortality Outcome | | Our Full 30-day Mortality Model | | | | | | | |
|---|---|---|---|---|---|---|---|---|---|
| | | **Outcome** | | | | **No Outcome** | | | |
| | | Lower Risk | Moderate Risk | High Risk | Extreme Risk | Lower Risk | Moderate Risk | High Risk | Extreme Risk |
| **VACO Index** | Lower Risk | <11* | | 126 | | 14101 | | 5535 | |
| | Moderate Risk | | <11* | | | | 2340 | | |
| | High Risk | | 47 | 26 | | | | 2381 | |
| | Extreme Risk | | | | 314 | | 10564 | | 3985 |
| | | | **Net correct reclassification:** | | 79 | | **Net correct reclassification:** | | 5029 |

| Legend |
|---|
| Correct Reclassification |
| Incorrect Reclassification |

**Absolute NRI:** +13%

* Data from Centers for Medicare & Medicaid Services (CMS) were combined with other data sources and masked in accordance with CMS cell size suppression policy (https://resdac.org/articles/cms-cell-size-suppression-policy)

Outcome is 30-day all-cause mortality. Risk groups for VACO index are based on published guidelines** and defined as: Lower Risk = lower 50% of scores, Moderate Risk = 4th sextile of scores, High Risk = 5th sextile of scores, and Extreme Risk = upper sextile of scores. Risk groups for our Mortality Model are defined similarly based on sextiles of the predicted risk in the holdout data set. nri

**MDCalc [Internet]. [cited 2023 Nov 3]. Veterans Health Administration COVID-19 (VACO) Index for COVID-19 Mortality. Available from: https://www.mdcalc.com/calc/10346/veterans-health-administration-covid-19-vaco-index-covid-19-mortality

#### S7b. Net Reclassification Index (NRI) of the 30-day all-cause mortality compared with the recalibrated CAN Score

| | | Our Full 30-day Mortality Model | | | | | | | |
|---|---|---|---|---|---|---|---|---|---|
| 30-day Mortality Outcome | | Outcome | | | | No Outcome | | | |
| | | Quartile - 1 | Quartile - 2 | Quartile - 3 | Quartile - 4 | Quartile - 1 | Quartile - 2 | Quartile - 3 | Quartile - 4 |
| Recalibrated CAN Score** | Quartile - 1 | <11* | | 56 | | 9206 | | 6424 | |
| | Quartile - 2 | | <11* | | | | 5539 | | |
| | Quartile - 3 | 11 | | 37 | | 3623 | | 7126 | |
| | Quartile - 4 | | | | 410 | | | | 6988 |
| | | | **Net correct reclassification:** | | 45 | | **Net correct reclassification:** | | -2801 |

| |
|---|
| Correct Reclassification |
| Incorrect Reclassification |

**Absolute NRI:** -7%

* Data from Centers for Medicare & Medicaid Services (CMS) were combined with other data sources and masked in accordance with CMS cell size suppression policy (https://resdac.org/articles/cms-cell-size-suppression-policy)

** the CAN score was recalibrated using Platt scaling

For calculated the NRI, we used the empirical quartiles of each model to group patients into quartiles of risk.

#### S8. SHAP Values of individual features in each outcome model

SHAP vaanxilues are derived from XGBoost model fitted in each ensemble model. Larger values indicate greater contributions to predicted probability of outcome. Table is sorted in descending order on the Composite outcome model.

| | Mean Absolute SHAP Value | | | Absolute SHAP Value Rank | | |
|---|---|---|---|---|---|---|
| **Feature** | **Composite outcome model** | **Hospitalization model** | **Mortality model** | **Composite outcome model** | **Hospitalization only model** | **Mortality model** |
| Care Assessment Needs Score | 0.603 | 0.554 | 1.326 | 1 | 1 | 1 |
| Charlson Comorbidity Index | 0.193 | 0.201 | 0.125 | 2 | 2 | 6 |
| 1a Facility Patient | 0.159 | 0.179 | 0.042 | 3 | 3 | 18 |
| Treatment - Paxlovid | 0.153 | 0.143 | 0.161 | 4 | 4 | 5 |
| Body Mass Index | 0.116 | 0.107 | 0.174 | 5 | 5 | 4 |
| Vaccination - Boosted | 0.105 | 0.099 | 0.100 | 6 | 6 | 7 |
| Age (years) | 0.092 | 0.078 | 0.197 | 7 | 7 | 2 |
| Days Since Last Vaccination | 0.059 | 0.061 | 0.054 | 8 | 8 | 13 |
| Treatment - Molnupiravir | 0.057 | 0.054 | 0.067 | 9 | 11 | 10 |
| FDAEUA01_age65 | 0.055 | 0.047 | 0.178 | 10 | 14 | 3 |
| FDAEUA08_CardiovascularDisease | 0.055 | 0.059 | 0.023 | 11 | 9 | 28 |
| CVD2yrs | 0.054 | 0.057 | 0.017 | 12 | 10 | 33 |
| Area Deprivation Index | 0.049 | 0.049 | 0.043 | 13 | 12 | 17 |
| FDAEUA18_other | 0.048 | 0.049 | 0.030 | 14 | 13 | 25 |
| FDAEUA06_immuno | 0.044 | 0.039 | 0.035 | 15 | 15 | 23 |
| Community Living Center Resident | 0.037 | 0.034 | 0.038 | 16 | 17 | 20 |
| Region - Midwest | 0.033 | 0.035 | 0.009 | 17 | 16 | 48 |
| Vaccinated | 0.029 | 0.015 | 0.084 | 18 | 31 | 8 |
| HTN2yrs | 0.028 | 0.029 | 0.009 | 19 | 19 | 47 |
| Smoking Status - Current | 0.025 | 0.015 | 0.058 | 20 | 29 | 12 |
| Region - South | 0.025 | 0.031 | 0.003 | 21 | 18 | 66 |
| DrugDep2yrs | 0.024 | 0.025 | 0.003 | 22 | 21 | 65 |
| PAD2yrs | 0.024 | 0.026 | 0.007 | 23 | 20 | 54 |
| Bipolar2yrs | 0.023 | 0.021 | 0.002 | 24 | 23 | 72 |
| HeartFailure2yrs | 0.023 | 0.016 | 0.048 | 25 | 28 | 16 |
| CANMort1yr30d_Unknown | 0.020 | 0.018 | 0.050 | 26 | 26 | 15 |
| Urban Resident | 0.020 | 0.023 | 0.002 | 27 | 22 | 70 |
| FDAEUA04_CKD | 0.018 | 0.010 | 0.014 | 28 | 43 | 35 |
| PTSD2yrs | 0.017 | 0.020 | 0.008 | 29 | 24 | 49 |
| FDAEUA02_bmi25 | 0.016 | 0.013 | 0.041 | 30 | 35 | 19 |
| Smoking_Unknown | 0.015 | 0.014 | 0.007 | 31 | 32 | 55 |
| Asthma2yrs | 0.015 | 0.013 | 0.026 | 32 | 36 | 26 |
| Immunocompromised | 0.014 | 0.011 | 0.037 | 33 | 41 | 21 |
| IscStroke2yrs | 0.014 | 0.014 | 0.000 | 34 | 33 | 90 |
| DiabetesOther2yrs | 0.013 | 0.014 | 0.001 | 35 | 34 | 75 |
| CHF2yrs | 0.013 | 0.015 | 0.012 | 36 | 30 | 38 |
| DiabetesWComp2yrs | 0.012 | 0.011 | 0.014 | 37 | 39 | 36 |
| FDAEUA17_SubstanceUseDisorder | 0.011 | 0.010 | 0.010 | 38 | 42 | 45 |
| DiabetesTypeTwo2yrs | 0.011 | 0.007 | 0.005 | 39 | 50 | 59 |
| Cancer2yrs | 0.011 | 0.007 | 0.034 | 40 | 49 | 24 |
| FDAEUA12_stroke | 0.011 | 0.012 | 0.000 | 41 | 37 | 87 |
| AlcDep2yrs | 0.011 | 0.009 | 0.009 | 42 | 45 | 46 |
| Parkinsons2yrs | 0.010 | 0.006 | 0.023 | 43 | 52 | 29 |
| FDAEUA07_addimmuno | 0.010 | 0.011 | 0.008 | 44 | 40 | 52 |
| Cirrhosis2yrs | 0.010 | 0.007 | 0.036 | 45 | 48 | 22 |
| MetasTumor2yrs | 0.009 | 0.001 | 0.059 | 46 | 90 | 11 |
| BloodTransfusion2yrs | 0.009 | 0.008 | 0.011 | 47 | 47 | 40 |
| FDAEUA15_mental | 0.009 | 0.018 | 0.050 | 48 | 25 | 14 |
| Ethnicity - Hispanic or Latino | 0.008 | 0.009 | 0.000 | 49 | 44 | 88 |
| MDD2yrs | 0.008 | 0.012 | 0.010 | 50 | 38 | 44 |
| Treatment - Monoclonal | 0.008 | 0.005 | 0.020 | 51 | 55 | 31 |
| FDAEUA13_liver | 0.008 | 0.009 | 0.001 | 52 | 46 | 86 |
| Race – Black or African American | 0.007 | 0.016 | 0.075 | 53 | 27 | 9 |

| | | | | | | |
|---|---|---|---|---|---|---|
| Region - West | 0.007 | 0.007 | 0.022 | 54 | 51 | 30 |
| Nephrosis2yrs | 0.007 | 0.005 | 0.001 | 55 | 57 | 76 |
| DiabetesWoComp2yrs | 0.007 | 0.003 | 0.011 | 56 | 64 | 42 |
| HeartDisease2yrs | 0.006 | 0.006 | 0.006 | 57 | 53 | 57 |
| CKF2yrs | 0.006 | 0.006 | 0.003 | 58 | 54 | 64 |
| COPD2yrs | 0.006 | 0.004 | 0.012 | 59 | 60 | 39 |
| Schizophrenia2yrs | 0.005 | 0.004 | 0.004 | 60 | 59 | 62 |
| Liver2yrs | 0.005 | 0.003 | 0.002 | 61 | 67 | 68 |
| Vaccination - Primary Series | 0.005 | 0.002 | 0.017 | 62 | 76 | 34 |
| Male | 0.005 | 0.004 | 0.006 | 63 | 62 | 58 |
| PulmHD2yrs | 0.005 | 0.003 | 0.008 | 64 | 68 | 51 |
| Epilepsy2yrs | 0.004 | 0.004 | 0.002 | 65 | 61 | 69 |
| PulmonaryFibrosis2yrs | 0.004 | 0.002 | 0.007 | 66 | 74 | 56 |
| DiabetesAny2yrs | 0.004 | 0.003 | 0.001 | 67 | 69 | 81 |
| HIV2yrs | 0.004 | 0.005 | 0.000 | 68 | 58 | 92 |
| CKD2yrs | 0.004 | 0.003 | 0.025 | 69 | 65 | 27 |
| ActMI2yrs | 0.003 | 0.005 | 0.002 | 70 | 56 | 73 |
| ChrLungDisease2yrs | 0.003 | 0.002 | 0.008 | 71 | 73 | 50 |
| Ethnicity_Unknown | 0.003 | 0.002 | 0.001 | 72 | 80 | 83 |
| ChrNeuroMuscDisease2yrs | 0.003 | 0.003 | 0.010 | 73 | 66 | 43 |
| Dialysis2yrs | 0.002 | 0.002 | 0.011 | 74 | 77 | 41 |
| MS2yrs | 0.002 | 0.002 | 0.001 | 75 | 79 | 80 |
| CAHD2yrs | 0.002 | 0.001 | 0.014 | 76 | 82 | 37 |
| Smoking Status - Former | 0.002 | 0.001 | 0.019 | 77 | 81 | 32 |
| FDAEUA09_lung | 0.002 | 0.002 | 0.008 | 78 | 78 | 53 |
| MildLD2yrs | 0.002 | 0.003 | 0.001 | 79 | 70 | 78 |
| Cerebrovasc2yrs | 0.002 | 0.004 | 0.003 | 80 | 63 | 67 |
| Race_Unknown | 0.002 | 0.003 | 0.003 | 81 | 71 | 63 |
| Emphysema2yrs | 0.002 | 0.002 | 0.004 | 82 | 72 | 61 |
| Race - Asian | 0.001 | 0.001 | 0.000 | 83 | 83 | 93 |
| Cardiomyopathy2yrs | 0.001 | 0.001 | 0.001 | 84 | 88 | 82 |
| ADI_NATRANK_avg_Unknown | 0.001 | 0.001 | 0.000 | 85 | 85 | 95 |
| Race - AI/AN | 0.001 | 0.001 | 0.001 | 86 | 87 | 84 |
| HemStroke2yrs | 0.001 | 0.000 | 0.001 | 87 | 91 | 85 |
| DiabetesTypeOne2yrs | 0.001 | 0.001 | 0.000 | 88 | 86 | 96 |
| OtherHD2yrs | 0.001 | 0.001 | 0.001 | 89 | 89 | 77 |
| Rural_Unknown | 0.000 | 0.001 | 0.000 | 90 | 84 | 94 |
| DialysisAtIndex | 0.000 | 0.000 | 0.000 | 91 | 92 | 89 |
| ChrRhHD2yrs | 0.000 | 0.000 | 0.000 | 92 | 96 | 91 |
| Race - Native Hawaiian/API | 0.000 | 0.000 | 0.000 | 93 | 94 | 97 |
| ChrHep2yrs | 0.000 | 0.000 | 0.001 | 94 | 97 | 74 |
| ModSevLD2yrs | 0.000 | 0.000 | 0.002 | 95 | 93 | 71 |
| Thalassemia2yrs | 0.000 | 0.000 | 0.000 | 96 | 95 | 98 |
| BMIAtIndex_Unknown | 0.000 | 0.002 | 0.004 | 97 | 75 | 60 |
| SickleCell2yrs | 0.000 | 0.000 | 0.001 | 98 | 98 | 79 |
| AgeAtIndexDate_Unknown | 0.000 | 0.000 | 0.000 | 99 | 99 | 99 |

#### S9. SHAP Values of individual features in each outcome model

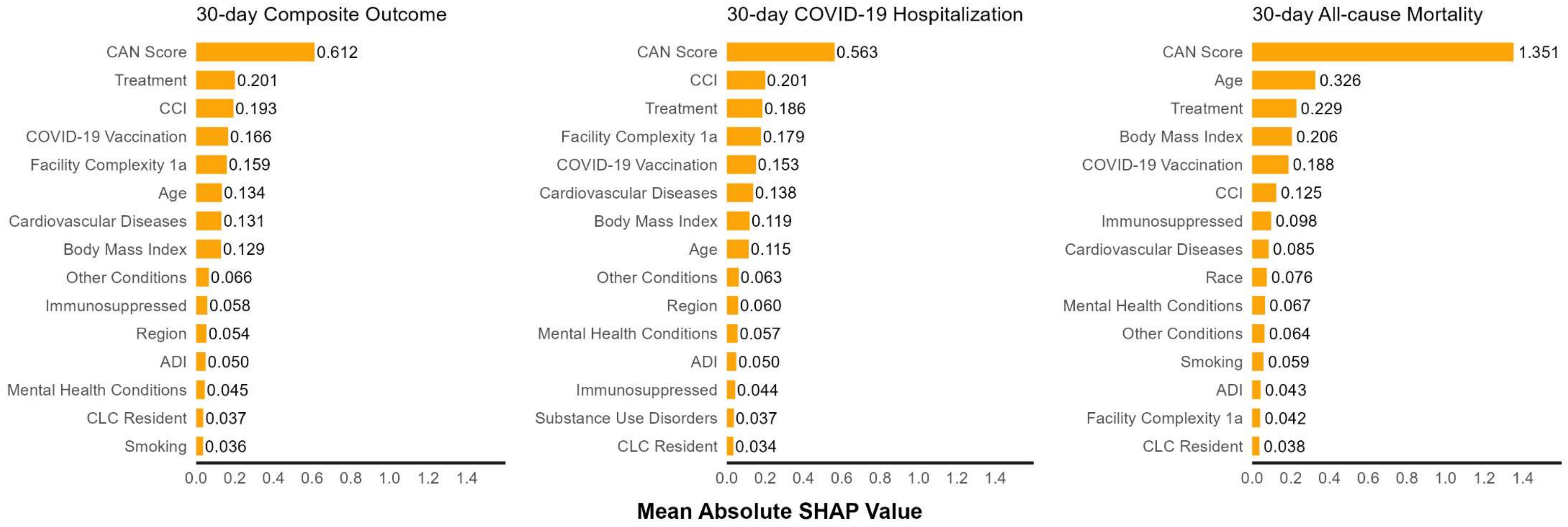

ADI: Area Deprivation Index
CCI: Charlson Comorbidity Index
CAN score: Care Assessment Needs score
CLC: community living center

Treatment SHAP value estimate includes indicators for receipt of outpatient nirmatrelvir-ritonavir, molnupiravir, or anti-SARS-CoV-2 monoclonal antibody treatment. Age includes age at index and indicator for being >65 years old. SHAP value estimates for cardiovascular diseases, immunocompromise, other diseases, substance use disorders, and mental health conditions, include SHAP values for all indicators within those domains (see supplement for list of indicators used in prediction).

S10. Predictors selected for parsimonious models

| No. | Variable Domain | Composite Model | Hospitalization Model | Mortality Model |
|---|---|---|---|---|
| 1 | Charlson Comorbidity Index | X | X | X |
| 2 | Age (years) | X | X | X |
| 3 | Age ≥ 65 years indicator | X | X | X |
| 4 | Paxlovid Treatment | X | X | X |
| 5 | Molnupiravir Treatment | X | X | X |
| 6 | Any Monoclonal Treatment | X | X | X |
| 7 | Vaccination Status | X | X | X |
| 8 | Days Since Last Vaccination | X | X | X |
| 9 | Body Mass Index | X | X | X |
| 10 | Overweight (body mass index ≥ 25) | X | X | X |
| 11 | Cardiovascular Disease | X | X | X |
| 12 | Immunosuppressive Condition – composite (any) | X | X | X |
| 13 | Region | X | X | X |
| 14 | Smoking Status | X | X | X |
| 15 | Race | | | X |
| 16 | Mental Health Conditions (any) | | | X |
| 17 | Heart Failure | | | X |
| 18 | Cirrhosis | | | X |
| 19 | Metastatic Tumor | | | X |

#### S11. Net reclassification index (NRI) for parsimonious models compared with fully fitted models.

| Full Model | | Parsimonious Model: Outcome Quartile-1 | Outcome Quartile-2 | Outcome Quartile-3 | Outcome Quartile-4 | No Outcome Quartile-1 | No Outcome Quartile-2 | No Outcome Quartile-3 | No Outcome Quartile-4 |
|---|---|---|---|---|---|---|---|---|---|
| 30-day Composite | Quartile-1 | 80 | | 253 | | 7719 | | 6009 | |
| | Quartile-2 | | 137 | | | | 5606 | | |
| | Quartile-3 | 502 | | 345 | | 5401 | | 5459 | |
| | Quartile-4 | | | | 1981 | | | | 5934 |
| 30-day Hospitalization | Quartile-1 | 80 | | 235 | | 7680 | | 6147 | |
| | Quartile-2 | | 138 | | | | 5531 | | |
| | Quartile-3 | 477 | | 312 | | 5543 | | 5415 | |
| | Quartile-4 | | | | 1676 | | | | 6192 |
| 30-day Mortality | Quartile-1 | <11* | | 16 | | 7879 | | 6022 | |
| | Quartile-2 | | <11* | | | | 5757 | | |
| | Quartile-3 | 37 | | 31 | | 5769 | | 5820 | |
| | Quartile-4 | | | | 430 | | | | 7659 |

| Legend |
|---|
| Correct Reclassification |
| Incorrect Reclassification |

* Data from Centers for Medicare & Medicaid Services (CMS) were combined with other data sources and masked in accordance with CMS cell size suppression policy (https://resdac.org/articles/cms-cell-size-suppression-policy)

Absolute NRI: Parsimonious composite model = -2.2.%; Parsimonious hospitalization model = -2.1%; Parsimonious mortality model = -0.7%
